# Dietary patterns and predictors of food insecurity and poor diet among children less than 5 years in the arid and semi-arid region of Kenya: a longitudinal study

**DOI:** 10.1101/2025.07.02.25330725

**Authors:** Chessa K Lutter, Linda Adair, Valerie L Flax, Faith Thuita, Estelle Sidze, Dickson Amugsi, Bonventure Mwangi, Albert Webale, Esther Anono, Hazel Odhiambo, Elizabeth Kimani-Murage, Calistus Wilunda

**Author notes:** Corresponding author Calistus Wilunda, African Population and Health Research Center, APHRC Campus, Kitisuru Nairobi, Kenya. Chessa Lutter is now located in Bethesda, MD USA.

## Abstract

We examined diet among children 6-23 and 24-59 months participating in a 2-year longitudinal study with six waves of data collection in Samburu and Turkana counties, Kenya. Information on child feeding practices was collected using methods recommended by the World Health Organization and UNICEF. Grains, roots, and tubers were nearly universally consumed. Dairy was the dominant animal source food (ASF), though declined sharply from Wave 1-5 among children 6-23 months and remained low across waves among children 24-59 months. The proportion of children who consumed fruits and vegetables, legumes and nuts, and eggs was low across waves. About half of children 6-23 months consumed sweet drinks at Wave 1 and consumption increased thereafter in both counties. Among children 24-59 months, nearly 90% of children in Samburu and about 65% in Turkana consumed a sweet drink across waves. Across all survey waves, only 11% and 5% of observations included foods from four or more food groups (representing minimum dietary diversity) in Samburu and Turkana, respectively. The mean number of food groups consumed ranged between 1.5 and 2.5, even among food secure households, though children in food secure households consumed about 0.5 more food groups. Children were more likely to consume dairy or any other ASF compared with their mothers, suggesting that mothers prioritize the nutritional needs of children over their own. The high prevalence of sweet drink consumption is mainly from sweetened tea, which is most likely given as a substitute for milk when milk is unavailable in pastoral and agropastoral households.

## INTRODUCTION

Children less than 5 years of age are the most affected by poor nutrition, including global acute malnutrition (GAM) (Black et al., 2013). To support rapid growth, young children require foods that are nutrient rich, especially animal source foods (ASFs), compared to older children and adults (L. L. Iannotti, 2018; Lutter, GrummerlStrawn, & Rogers, 2021).

Therefore, they are more affected by household food insecurity. Throughout low- and middle-income countries, the diets of young children vary within and among countries and are generally poor (UNICEF, 2021), particularly in Samburu and Turkana counties in Northwest, Kenya (Kenya National Bureau of Statistics & ICF, 2023).

Characterized by a hot and dry climate, low and erratic rainfall patterns, recurrent drought, conflict, and other shocks (Samburu County Government, 2023; Turkana County Government, 2023), households in Samburu and Turkana are affected by deep-rooted poverty, lack of access to basic services, poor food availability, food insecurity, low levels of formal education, and migration. Not surprisingly, levels of acute and chronic malnutrition are persistently high and feeding practices among children less than 5 years are far from optimal (Samburu County Department of Health, 2023; Turkana County Department of Health, 2023). According to the 2022 Demographic and Health Survey (Kenya National Bureau of Statistics & ICF, 2023), 30% and 32% of children under 5 were acutely malnourished in Samburu and Turkana, respectively, well above the Sphere emergency threshold (Sphere, 2018).

Samburu and Turkana counties are inhabited by pastoralists and agropastoralists, but there has been a movement toward settlement in urban and semi-urban settings. In Turkana, there is also a community of fisherfolk along Lake Turkana. Traditionally, animal milk has supported child nutrition in pastoral and agropastoral communities (L. Iannotti & Lesorogol, 2014). However, in addition to increasingly erratic climatic patterns, the privatization of land tenure, land conservation, and encroachment of urban settlements has restricted livestock movements and increased resource competition. Economic development and urbanization in the counties have had both positive and negative effects on child nutrition (Young & Marshak, 2018). Access to health care, water and sanitation infrastructure, and economic opportunities for diversification of household income should have positive effects unless the quality and accessibility of health care is poor, as is the case in these counties. At the same time, the commercialization of livestock and urbanization have undermined traditional kinship systems and social support institutions, negatively impacting child nutrition.

In both counties, the diets of children under 2 years are far from optimal. In 2018 in Samburu, a Knowledge, Attitudes, Beliefs, and Practices (KABP) survey showed that only 48% of infants 6–8 months of age received complementary foods (Ochola, 2018). Among children 6– 23 months of age, only 36% met the minimum meal frequency and 60% attained the minimum dietary diversity. Dairy was the most consumed (88% of children), followed by cereals (84% of children). Around 50% of children consumed roots or tubers; legumes; or beans, lentils, or nuts. A dark green vegetable or a vitamin A rich fruit was consumed by about 42% of children and eggs by 36%. Fish was consumed by less than 2% of children. Slightly over 40% of children consumed meat and 37% organ meat. The reported high consumption of meat and organ meat is difficult to reconcile with other information in the report stating that only 21% of children received an iron-rich food as the numbers should be similar.

Compared to Samburu, complementary feeding practices in Turkana are worse (Ochola, 2017). A similar KABP survey showed that only 36% of infants 6–8 months of age received complementary foods. Among children 6–23 months, only 36% attained the minimum meal frequency and 46% attained the minimum dietary diversity. Iron-rich foods were consumed by nearly half (48%) of children. As in Samburu, cereals were most consumed (80% of children) followed by dairy (70% of children) and beans, lentils, or nuts (57% of children). Around one-third of children consumed root or tubers, dark green vegetables. A total of 37% of children consumed meat, 33% consumed organ meat, and 15% consumed eggs.

As part of the USAID-funded Nawiri project, a flagship initiative to sustainably reduce levels of acute malnutrition among young children in Kenya’s arid and semi-arid lands, we conducted a 2-year longitudinal study. The purpose was to understand and map how a variety of immediate, underlying, basic or systemic factors influence global acute malnutrition (GAM) among children less than 5 years over time and by livelihood and identify and prioritize opportunities to achieve sustained reductions in GAM. Previously published papers have summarized the protocol of the study and reported on the factors associated with acute malnutrition (Amugsi et al., 2025; Wilunda et al., 2024). In this paper, we report results related to child diet. Specifically, we ask four questions: 1) how does children’s consumption of different food groups vary over the 2-year period? 2) how does the proportion of children consuming foods from specific food groups vary by the number of food groups consumed? 3) what are the factors associated with food insecurity and child diet? and 4) how do dietary patterns of mothers/caregivers differ from those of children?

## KEY MESSAGES

- Young children in Samburu and Turkana counties are receiving a diet that is far from satisfying their nutritional needs for growth and development.
- Over the 2-year study period, consumption of animal source foods decreased to less than 1% in both counties, while that of sweet beverages increased to almost 90% in Samburu and 65% in Turkana.
- Across all six survey waves, only 11% of observations from Samburu and 5% of observations from Turkana included foods from four or more food groups.
- In both counties, the mean number of food groups consumed ranged between 1.5 and 2.5 across study waves, well below the minimum dietary diversity cutoff, even among food secure households.
- Consumption of ASFs in both counties was significantly negatively associated with low wealth index tertile and severe food insecurity, and positively associated with household ownership of chickens or a TV or radio.

## METHODS

### Study design

Details of the design, sample size estimation, and sampling strategy are described elsewhere (Wilunda et al., 2024). In brief, a multistage sampling strategy was used so that findings were representative at both the county and sub-county levels. The population was stratified according to survey zones, as defined by respective county governments. Within each survey zone, villages were randomly selected for inclusion. Locally hired field enumerators conducted a household listing in the selected villages to create a sample frame of households with children under 3 years of age. Households were then randomly selected for participation in the study. In Samburu, data were collected every 4 months over 2 years, from June/July 2021 to August 2023 for a total of six distinct waves of data collection. According to the Kenyan National Drought Management Authority (NDMA), the early warning classification was at the normal level only during Wave 1 (enrollment). In Turkana, data were also collected every 4 months over 2 years from May–June 2021 to August – September 2023. Except for Wave 1, throughout the rest of the study, early warning classifications were at the alert, alarm, emergency, or recovery levels, according to the NDMA.

Stratification by livelihood was done through post-stratification analysis. Data were analyzed for three livelihoods in Samburu, pastoral, agropastoral, and urban/peri-urban, and four livelihoods in Turkana, pastoral, agropastoral, fisherfolk, and urban/peri-urban. In Samburu, 586 households were included at Wave 1 and 544 were surveyed at Wave 6 (93% response rate). In Turkana, 1,211 households were included at Wave 1 and 1,156 were surveyed at Wave 6 (96% response rate). For analysis of child diet, we only include children ages 6-59 months with complete diet and covariate data (465 and 999 children in Samburu and Turkana countries, respectively, at Wave 1). Allowing for repeated measures, across the six waves dietary data included 3,163 observations in Samburu and 6,447 observations in Turkana.

### Data collection

At the individual level, we collected sociodemographic data from the caregiver of the index child on age, education, marital status, number of children, sex of the household head, household food insecurity, and other household sociodemographic characteristics.

Information on child feeding practices the day prior to the survey, including breastfeeding and child diet (for all children less than 5 years), was collected from the caregiver using a methodology recommended by the World Health Organization and UNICEF (World Health Organization & UNICEF, 2021). First, the caregiver was asked about plain water, infant formula, milks, juice/juice drinks, sodas other sweet drinks, clear broth, yogurt, thin porridge, other liquids (e.g., ORS type fluids), and other liquids (tea, coffee). Second, they were asked to recall everything the child ate yesterday as well as ingredients in mixed dishes. Third, the interviewer underlined foods in the list provided by the WHO/UNICEF questionnaire and read those back to get a yes-no answer to food items fed yesterday. Fourth, there was an additional question about semisolid or soft foods and these were added if not already on the list. Lastly, there was a list of fortified foods.

### Analysis by study question

#### How does children’s consumption of different food groups vary over the 2-year period?

Across the six study waves, we estimated the unweighted proportion of children consuming foods from selected food groups: animal source foods (ASFs), fruits and vegetables, legumes and nuts, and sweet beverages, in addition to breast milk. We chose these foods because they represent the main sources of micronutrients and protein in the diet. We examined sweetened beverages as a potentially detrimental food group. We did not separately include grains, since they were consumed by nearly all children. Further, the generally low consumption of the less aggregated groups (e.g., eggs) gave us enough observations for meaningful analysis.

The analysis was stratified by child age (6-23 months and 24-59 months to account for changing breastfeeding practices among older children) and county.

#### How does the proportion of children consuming foods from specific food groups vary by the number of food groups consumed?

For this analysis, to be consistent with the food groups recommended by WHO and UNICEF for infant and young child feeding (World Health Organization & UNICEF, 2021), we summed the seven food groups (grains, roots, and tubers (including thin gruels); dairy; vitamin A rich fruits and vegetables; legumes and nuts; other fruits and vegetables; meat and fish; and eggs) to provide a score, which ranged from one to seven (consumed foods from one group to all seven groups). To better understand which food groups typically comprised each level of the score, we examined the frequency of consumption of each food group at each level of the food group summary score. Children with zero food groups were either exclusively breastfed or were given other liquids, which included teas, honey, or broth, the composition of which was not recorded. The latter group of children was excluded from the analysis.

#### What are the factors associated with food insecurity and child diet?

For the analysis of the factors associated with food insecurity and child diet, we used sections of the conceptual framework for acute malnutrition in Africa’s drylands (Young & Marshak, 2018). This framework illustrates how various basic and underlying factors affect child dietary intake, which is an intermediate factor in the framework (Young & Marshak, 2018). To represent child dietary intake, we used consumption from two key food groups: 1) all ASFs (meat, fish, eggs, and dairy products) and 2) all types of fruits and vegetables. We used these two groups because bivariate analyses suggested their importance in relation to wasting, defined as weight-for height Z-score <-2 SD), in the sample. We used the Food Insecurity Experience Scale (FIES) (Ballard, Kepple, & Cafiero, 2013) to obtain food insecurity prevalence rates. We then categorized the FIES data as a binary variable to represent severe food insecurity, which is an underlying factor in the framework.

We used mother/caregiver alcohol use (yes, no) as a proxy for inadequate caregiving practices, another underlying variable. The basic variables we included were mother/caregiver characteristics, including education (i.e., any formal education or not), marital status (unmarried, married to one wife, or married with co-wives), and whether the caregiver was the biological mother of the child. The household characteristics included were female-headed household; mother or caregiver number of children; household wealth index tertiles (highest, middle, lowest); ownership of poultry; ownership of a radio or a TV; and whether the household had experienced climate, economic, biological, or conflict shocks since the last visit, expressed as total number of shocks (0–4). To take advantage of the longitudinal data, we used generalized estimating equations for each outcome across all six survey waves and ran five sets of models. An initial set of models identified bivariate associations, which we then used to inform multivariable models. All multivariable models included survey zone, livelihood, survey wave, and child’s age and sex.

#### How do dietary patterns of mothers/caregivers differ from those of children?

Choosing Wave 3 (which for Samburu was completed in April-May, 2022 and for Turkana May-June, 2022) we estimated the percentage of mothers/caregivers and children consuming fruits and vegetables, dairy, legumes and nuts, and ASFs. Since stratification by age revealed no differences in the observed relationships, we report results for the full 6–59-month sample. Assuming that food items are available in the household if mothers/caregivers are consuming them, we defined a “missed opportunity” when the mother/caregiver consumed, but the child did not consume those foods or vice versa.

### Ethics statement

We obtained ethical and research approvals and research permits from the Ethical and Scientific Review Committee (ESRC) of Amref Health Africa (Amref - ESRC P905/2020) and the National Commission for Science, Technology, and Innovation of Kenya, respectively. The institutional review boards at APHRC and RTI also signed a reliance agreement. We obtained informed consent from all participants during Wave 1 and reconfirmed it at Waves 2–6.

## RESULTS

### Descriptive characteristics

We summarize Wave 1 data on child, caregiver, household, and community characteristics for children with dietary data for all six waves, by county in **Table 1**. The unweighted estimates of the prevalence of wasting were 13.8% and 25.2% in Samburu and Turkana, respectively. Child sex ratios were similar in Samburu, whereas in Turkana there were more male than female children. The proportion of children aged 12–23-months was similar between the two counties, but there were more children in the 0–11-month age category in Samburu and more children in the 24–59-month age category in Turkana. Samburu had far more mothers/caregivers less than 25 years of age than Turkana, whereas in Turkana there were more mothers/caregivers 35 years and older. Marital status also differed between counties; there were fewer married women with co-wives and women who were not married in Samburu compared with Turkana. Although most mothers/caregivers did not have formal education, there were far more in this category in Turkana compared with Samburu.

**Table 1:** Child, caregiver, household, and community characteristics at Wave 1, by county.

|  | **Samburu (N=465)** | **Turkana (N=999)** |
| --- | --- | --- |
|  | **%** | **%** |
| ***Child Characteristics*** |  |  |
| Prevalence of wasting | 13.8<sup>a</sup> | 25.2<sup>b</sup> |
| Sex |  |  |
| Male | 50.3 | 54.8 |
| Female | 49.7 | 45.2 |
| Age (months) |  |  |
| 0-11 | 39.1 | 24.4 |
| 12-23 | 45.2 | 41.8 |
| 24-59 | 15.7 | 33.7 |
| ***Caregiver/Mother Characteristics*** |  |  |
| Age (years) |  |  |
| <25 | 39.1 | 3.6 |
| 25-34 | 45.2 | 63.8 |
| 35 and above | 15.7 | 32.6 |
| Marital status |  |  |
| Married only wife | 58.1 | 46.9 |
| Unmarried | 14.8 | 19.9 |
| Married co-wife | 27.1 | 33.1 |
| Caregiver number of children |  |  |
| Less than 2 | 33.0 | 25.9 |
| 3-5 | 37.3 | 49.6 |
| 6 or more | 29.7 | 24.4 |
| Highest level of education |  |  |
| Formal education | 33.3 | 13.6 |
| No formal education | 66.7 | 86.4 |
| ***Household Characteristics*** |  |  |
| Age of the household head (years) |  |  |
| <25 | 11.3 | 10.8 |
| 25-34 | 36.3 | 34.5 |
| 35 and above | 52.4 | 54.6 |
| Sex of household head |  |  |
| Male | 82.8 | 55.8 |
| Female | 17.2 | 44.2 |
| Ownership of poultry and TV or radio |  |  |
| Household owns poultry | 89.3 | 64.7 |
| Household owns TV or radio | 41.1 | 13.4 |
| Household size |  |  |
| < 4 | 13.6 | 11.8 |
| 4 - 6 | 52.7 | 49.8 |
| 7+ | 33.8 | 38.4 |
| Number of shocks<sup>c</sup> |  |  |
| 1 | 6.1 | 6.9 |
| 2 | 29.8 | 25.5 |
| 3 | 29.8 | 39.2 |
| 4 | 26.4 | 26.1 |
| ***Community Variables*** |  |  |
| Livelihood |  |  |
| Urban/peri-urban | 18.7 | 42.7 |
| Pastoral | 59.1 | 36.7 |
| Agropastoral | 22.2 | 13.0 |
| Fisherfolk | n.a | 7.5 |
| Survey zone |  |  |
| Central | 52.0 | 28.4 |
| North | 23.4 | 12.3 |
| East | 24.5 | n.a |
| South | n.a | 33.1 |
| West | n.a | 26.1 |

<sup>a</sup> Prevalence of wasting based on a sample of 435 children
<sup>b</sup> Prevalence of wasting based on a sample of 939 children
<sup>c</sup> Categorized as climatic (e.g, excessive rain and drought), economic (e.g, loss of livelihood and increased food prices), biological (e.g, livestock, crop, or human disease outbreak), or conflict (e.g., destruction of assets and theft of livestock).

With respect to household characteristics, far more households in Samburu were headed by a male compared with Turkana. Households in Samburu were also wealthier as assessed by ownership of poultry or a TV or radio. In both counties, households were exposed to between two and four shocks. In Turkana, the urban/peri-urban livelihood was dominant, whereas in Samburu pastoralism was dominant (**Table 1**).

### Children’s consumption of different food groups over the 2-year period

Child feeding practices were poor in both counties (**Figure 1**). The proportion of children who consumed fruits and vegetables and legumes and nuts were low in both counties and age groups across survey waves. Dairy was the dominant food in the ASF category. Across survey waves, dairy consumption declined sharply among children 6-23 months and remained low among older children in both counties. About half of children in the younger age category consumed sweet drinks at Wave 1 and consumption increased thereafter in both counties. Among children 24-59 months, nearly 90% of children in Samburu and 65% in Turkana consumed a sweet drink across study waves.

**Figure 1.**
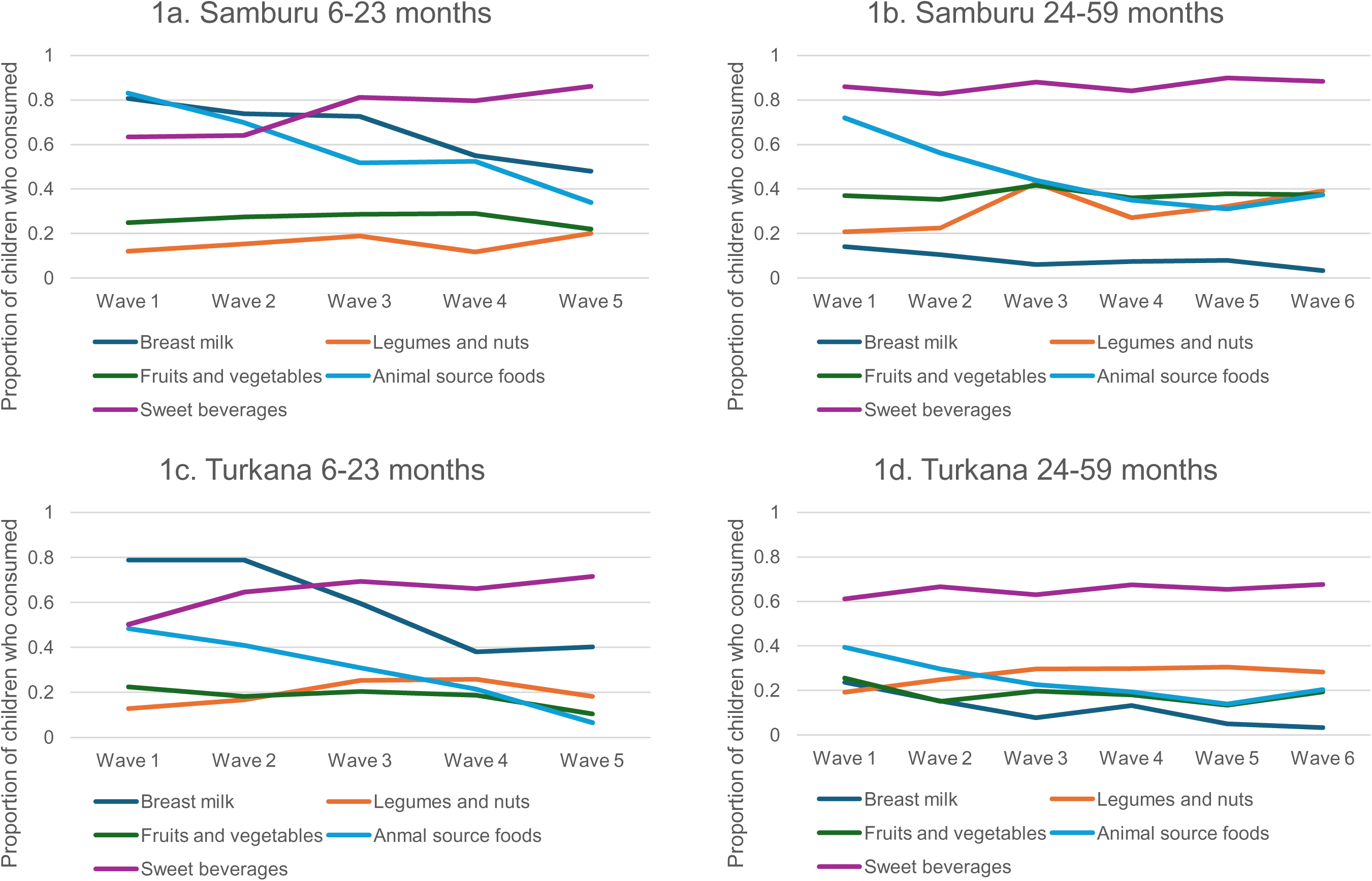
Pattern of children’s food group consumption over two years.

### Proportion of children consuming specific groups by number of food groups consumed

The proportion of children consuming foods from each of the seven food groups at each level of the food group is shown in **Table 2**. Across all survey waves, only 11% of observations from Samburu and 5% of observations from Turkana included foods from four or more food groups, indicating that they achieved minimum dietary diversity. In both counties, regardless of the number of food groups consumed, grains, roots, and tubers were nearly universally consumed. In Samburu, the four most common food groups were most often grains, roots, and tubers; dairy; legumes and nuts; and Vitamin A rich fruits and vegetables. Only 26% of the observations where foods from four groups were consumed included meat or fish. In Turkana, the four most common food groups were grains, roots, and tubers; dairy; vitamin A rich fruits and vegetables; and other fruits and vegetables. Only 53% of the observations when foods from four groups were consumed included meat or fish. Egg consumption was low in both counties. The table further shows very few observations (21 in Samburu and 16 in Turkana) where foods from six food groups were consumed and even fewer where foods from all seven food groups were consumed (three observations in Samburu and four in Turkana).

**Table 2.**
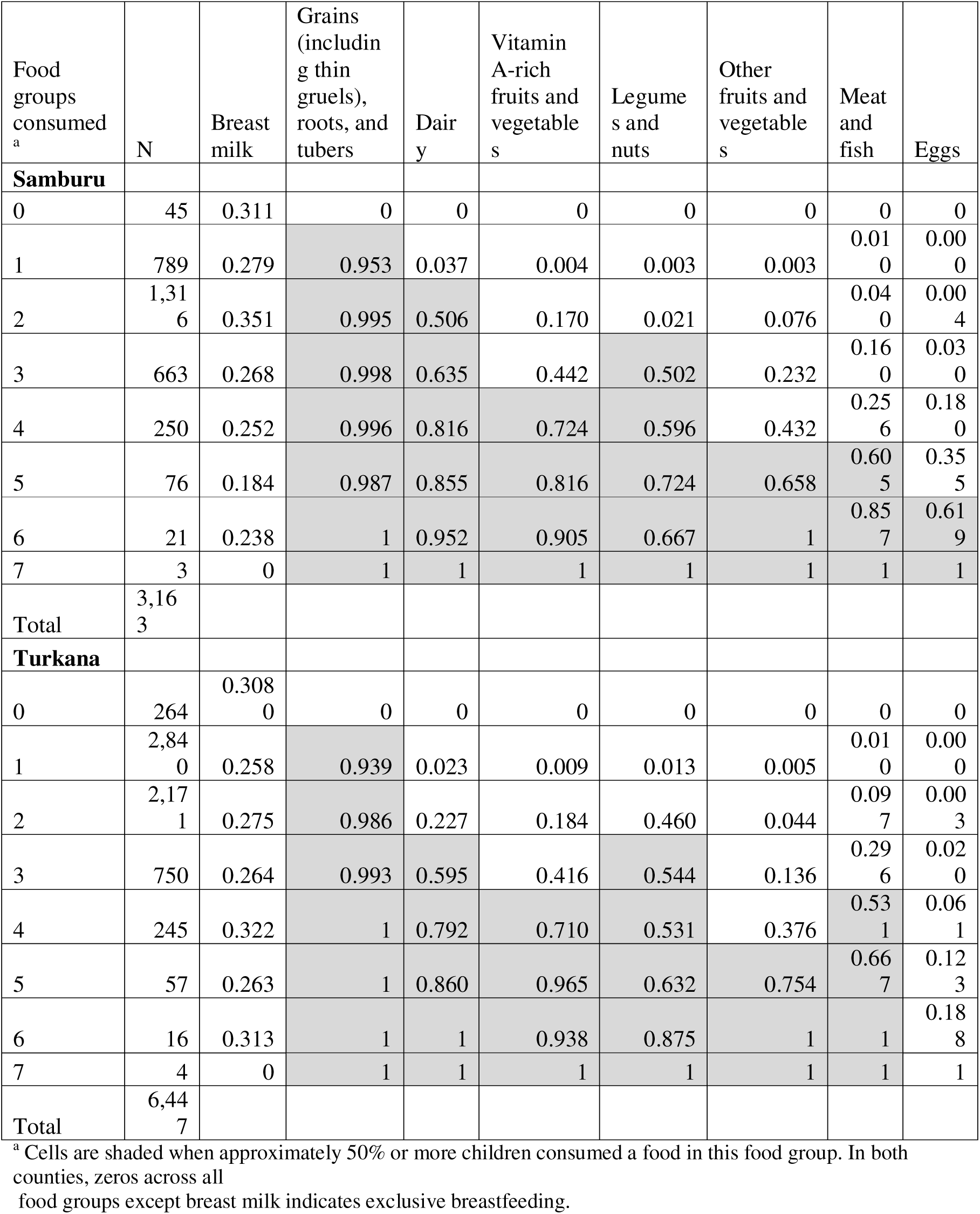
Proportion of children consuming each of the seven food groups in the number of food groups variable, by number of food groups consumed.

In both counties, the mean number of food groups consumed ranged between 1.5 and 2.5 across study waves, even among food secure households. Children in food secure households consumed about 0.5 more food groups than those from food insecure households (data not shown).

### Factors associated with food insecurity and child diet in Samburu

Results for factors associated with food insecurity and diet in Samburu are shown in Supplemental Table 1.

#### Model 1: Household food insecurity

On average across all waves, severe food insecurity was more likely if the mother/caregiver did not have any formal education, was unmarried, or if the child lived in a household in the middle or lower wealth index tertile, no poultry, or no TV or radio. Severe food insecurity was also more likely in households experiencing more economic, biological, and economic shocks. A lower likelihood of severe food insecurity was associated with living in the East zone and among agropastoralists.

#### Models 2 and 3: Basic or systemic factors related to child dietary adequacy

On average, across all survey waves, children were less likely to consume ASFs if their mother/caregiver lived in a household in the lowest wealth tertile or they were older than 2 years. Children in households that owned poultry were more likely to consume ASFs. When looking at waves, the odds of consuming ASFs was lower for all subsequent survey waves compared with Wave 1. Livelihood was not associated with child ASF consumption; however, children living in the East zone were more likely to consume ASFs. Breastfeeding status did not change any of these relationships.

When examining children’s consumption of all types of fruits and vegetables, children were less likely to consume such foods if their mother/caregiver lived in a household with a co-wife or the household was in the lowest or middle wealth index tertile. They were more likely to consume fruits and vegetables if they were older than two years or the caregiver had formal education. Children in the pastoral livelihood or in the East and North zones were less likely to consume fruits and vegetables.

#### Model 4: Severe food insecurity and dietary adequacy

Children were less likely to consume ASFs if their household had severe food insecurity. Food insecurity was not associated with fruits and vegetables consumption.

#### Model 5: Caregiver alcohol use related to dietary adequacy

Mother/caregiver alcohol use was not associated with children’s ASF or fruit and vegetable consumption.

### Factors associated with food insecurity and child diet in Turkana

Results for factors associated with food insecurity and diet in Turkana are shown in Supplemental Table 2.

#### Model 1: Basic or systemic factors related to household food insecurity

Severe food insecurity was less likely if the mother/caregiver had any formal education or the household owned poultry. Severe food insecurity was more likely among households where the caregiver had three or more children, the household belonged to the lowest or middle wealth index tertile, experienced two to four types of shock, was in the North, South, or West survey zones, was in the pastoral livelihood zone, and during Waves 3, 5, or 6, compared to Wave 1.

#### Models 2 and 3: Basic or systemic factors related to child dietary adequacy

Children were less likely to consume ASFs if they were older than two years, their caregiver had three or more children, the household was in the lowest or middle wealth index tertile, and during all survey waves compared to Wave 1. Children were more likely to consume ASFs if their mother/caregiver had any formal education, the household owned poultry, a TV or a radio, or the household reported their livelihood as fisherfolk. Children were less likely to consume fruits and vegetables if their mother/caregiver had three or more children, the household was in the lowest or middle wealth index tertile, in the North or West survey zones, were in the pastoral or fisherfolk livelihood compared to the urban/peri-urban livelihood, and during survey Waves 2 and 5 compared to Wave 1. Consumption of fruits and vegetables was more likely among female children, children with mothers/caregivers with any formal education or if the household owned a TV or radio.

#### Model 4: Household food security related to dietary adequacy

Children were less likely to consume ASFs or fruits and vegetables if their household had severe food insecurity.

#### Model 5: Mother/Caregiver alcohol use related to dietary adequacy

Mother/caregiver alcohol use was not associated with children’s ASF or fruit and vegetable consumption.

### Dietary patterns of mothers/caregivers compared to those of children

In both counties, children were more likely to consume dairy and any ASF compared with their mother/caregiver (**Table 3**). In Samburu, more mothers/caregivers consumed fruits and vegetables and legumes and nuts compared with children. However, in Turkana the reverse was true, and children were more likely to consume fruits and vegetables and legumes and nuts compared with their mother/caregiver.

**Table 3.** Concordance of mother/caregiver and child diets at Wave 3.

|  | Mothers/<br>caregivers who<br>consumed (%) | Children who<br>consumed (%) | "missed<br>opportunity" (%)<sup>a</sup> | Number<br>affected |
| --- | --- | --- | --- | --- |
| **Samburu (N = 522**<br>**mother/caregiver-child**<br>**pairs)** |  |  |  |  |
| Fruits and vegetables | 43.8 | 35.3 | 28.4 (child) | 65 |
| Dairy | 18.0 | 41.9 | 65.0<br>(mother/caregiver<br>) | 33 |
| Legumes and nuts | 44.8 | 31.0 | 35.7 (child) | 86 |
| Any animal source food | 32.0 | 47.7 | 25.7<br>(mother/caregiver<br>) | 43 |
| **Turkana (N=1,019 mother/**<br>**caregiver-child pairs)** |  |  |  |  |
| Fruits and vegetables | 19.8 | 20.2 | 27.5<br>(mother/caregiver<br>) | 56 |
| Dairy | 7.1 | 20.4 | 23.6<br>(mother/caregiver<br>) | 17 |
| Legumes and nuts | 28.0 | 38.4 | 29.6<br>(mother/caregiver<br>) | 151 |
| Any animal source food | 15.1 | 25.9 | 11.0<br>(mother/caregiver<br>) | 32 |

<sup>a</sup>Among caregivers who consumed food group, percentage of children who did not consume that group or vice versa.

## DISCUSSION

Similar to other studies that examined factors underlying child undernutrition (Victora et al., 2021 Mar), our study showed that poor child diets and household food insecurity were associated with poverty and household size. For example, the child’s mother/caregiver having no formal education, more than three children, consuming alcohol, or if the household was poor (assessed either as a function of wealth index tertile or household ownership of poultry, a TV, or a radio) were significant in many of the models. Like other studies, consumption of ASFs in both counties was significantly negatively associated with measures of wealth, such as belonging to a low wealth index quartile, and positively associated with household ownership of chickens or a TV or radio (Mussa et al., 2024).

Our study shows that young children in Samburu and Turkana counties are not receiving a diet that begins to satisfy their nutritional needs for growth and development. Additionally, it reveals that the traditional cultural practice of feeding young children on animal milk may be changing to the detriment of child nutrition and that within the same household dietary intake patterns differ between mothers/caregivers and young children (Mosites et al., 2016).

Over the 2-year study period, among children less than 3 years at baseline, consumption of ASFs decreased to less than 1% in both counties, while that of sweet beverages increased to almost 90% in Samburu and 65% in Turkana. Globally, the percentage of children 6-23 months consuming flesh foods ranged from 25% to 39% (UNICEF, 2021), illustrating the large gap between the diets in our study and that of young children worldwide, which are also not optimal. Traditionally, milk is given to young children in pastoralist societies and highly valued as a nutritious food even when pastoralism is no longer practiced (Young, 2021). When milk is not available, sweet teas are given as a substitute, which is reflected in the high consumption of sweet beverages.

Similar to many other countries, foods from the grains, roots, and tubers food category were by far the most frequently consumed (UNICEF, 2021). Such foods, while providing energy, are low in critical nutrients, such as iron and zinc, and have low protein quality (FAO., 2013). Many are also high in phytates that inhibit micronutrient absorption. Although grains, roots, and tubers were most consumed in both counties, county food group patterns diverged thereafter. In Samburu, dairy, primarily in the form of milk, was the next most common food group, followed by Vitamin A-rich fruits and vegetables, legumes and nuts, and other fruits and vegetables. In Turkana, other fruits and vegetables, dairy also primarily in the form of milk, Vitamin A-rich fruits and vegetables, and meat and fish were the next most important food groups. In both counties, eggs were rarely consumed, which is similar to the case in most low- and middle-income countries (L. L. Iannotti, Lutter, Bunn, & Stewart, 2014). What is surprising is how few food groups were consumed, ranging from an average of 1.5 to 2.5 across study waves, compared to global data that, again, do not reflect optimal young child diets (UNICEF, 2021). Consuming a diverse diet is important as it reduces the risk of micronutrient deficiencies and creates synergies that facilitate the absorption of nutrients (Lutter et al., 2021). The recently updated WHO guideline for complementary feeding recommends daily consumption of ASFs, fruits and vegetables, and pulses, nuts, and seeds to enhance dietary diversity (WHO, 2023). Grains, roots, and tubers are not included in this list, largely because dietary modeling exercises carried out during the development of the guideline revealed that including even small quantities of such foods increased the risk of diets not meeting dietary requirements. The fact that children are more likely to consume dairy and any ASFs compared to their mothers/caregivers suggests that mothers prioritize the nutritional needs of children over their own in conditions of scarcity (Atree, 2005). In Samburu, because mothers/caregivers were more likely to consume fruits and vegetables and legumes and nuts compared to children, promoting these foods, clearly available in the household, could be an intervention strategy to improve young children’s diets in the short term. This is not the case in Turkana, where children were more likely to consume foods from these groups than their mothers/caregivers.

In summary, our study showed extremely poor diets among children under 5 years in Samburu and Turkana. This is likely the result of multiple factors, including climate change, conflict in some areas of Turkana, changing patterns of land tenure, land conservation, and encroachment of urban settlements that have restricted livestock movements and increased resource competition. While evidence shows positive dietary and nutrition outcomes can be achieved through nutritionlspecific interventions, including nutritional counseling in the community and through health systems (Bhutta et al., 2013), except for counseling and support for breastfeeding, dietary interventions depend on household food security as well as caregiver/maternal knowledge about optimal young child feeding practices. There is also evidence to suggest that sustaining nutrition interventions is limited in the absence of external resources (Kim, Rogers, Coates, Gilligan, & Sarriot, 2013). Strengthening social safety nets so households can access adequate foods, including those needed for young children, is needed to address this dire situation in the short term. In the long term, given climate change and the rapidly changing socio-demographic context, addressing the underlying factors will require long-term investments to improve maternal education, and income.

## Data Availability

The data underlying this study will be made available to researchers through the Microdata Portal of APHRC (https://aphrc.org/microdata-portal/).

## Acknowledgments

None.

## Data sharing

The data underlying this study will be made available to researchers through the APHRC’s Microdata Portal (https://aphrc.org/microdata-portal/).

## Author contributions

CKL, VLF, FT, ES, and DA designed the study. CW, DA, BM, AW, FT, EA, and HO implemented fieldwork. CW, ES, FT, and EK-M provided study oversight. LA designed the analytic framework and analyzed the data. CKL prepared the first draft of the manuscript and incorporated comments from the co-authors. All authors contributed to the revision of the manuscript and approved the final version.

