## Supplemental Table 1 for "Dietary patterns and predictors of food insecurity and poor diet among children less than 5 years in the arid and semi-arid region of Kenya: a longitudinal study"

**Supplementa1 Table 1: Determinants of household food insecurity and child diet in Samburu**

|  | **Severe food insecurity** | **Child consumed**  **animal-source food** | **Child consumed**  **fruits or vegetables** |
| --- | --- | --- | --- |
|  | **aOR (95% CI)** | **aOR (95% CI)** | **aOR (95% CI)** |
| Number of children | **584** | **584** | **584** |
| Number of observations | **3163** | **3163** | **3163** |
| **Basic factors^a^** |  |  |  |
| Female child (ref. male child) | 1.02 (0.83 - 1.25) | 1.01 (0.83 - 1.23) | 1.17 (0.95 - 1.43) |
| Child aged ≥2 y (ref. <2 y) | 1.17 (0.95 - 1.45) | 0.63 (0.52 - 0.77)*** | 1.54 (1.23 - 1.93)*** |
| Caregiver educated (ref. not educated) | 0.68 (0.53 - 0.89)** | 1.25 (0.97 - 1.63) | 1.64 (1.27 - 2.12)*** |
| Caregiver age in y (ref. <25) |  |  |  |
| 25-34 | 1.01 (0.78 - 1.29) | 0.98 (0.77 - 1.24) | 0.80 (0.61 - 1.04) |
| 35 and above | 1.33 (0.96 - 1.84) | 0.84 (0.61 - 1.14) | 0.98 (0.71 - 1.36) |
| Caregiver marital status  (ref. married, only wife) |  |  |  |
| Unmarried | 1.58 (1.08 - 2.31)* | 0.99 (0.70 - 1.41) | 0.80 (0.55 - 1.14) |
| Married co-wife | 1.19 (0.92 - 1.53) | 1.19 (0.94 - 1.50) | 0.62 (0.48 - 0.82)*** |
| Household head is female (ref. male) | 1.07 (0.73 - 1.58) | 0.98 (0.69 - 1.41) | 1.28 (0.89 - 1.84) |
| Caregiver is child’s biological mother | 1.35 (0.86 - 2.12) | 1.08 (0.70 - 1.65) | 1.29 (0.84 - 1.98) |
| Caregiver number of living children (ref. 0-2) |  |  |  |
| 3-5 | 1.24 (0.96 - 1.61) | 0.94 (0.73 - 1.21) | 0.86 (0.65 - 1.13) |
| 6 and above | 1.32 (0.96 - 1.81) | 0.84 (0.62 - 1.13) | 0.89 (0.64 - 1.22) |
| Household owns poultry | 0.60 (0.45 - 0.80)*** | 1.62 (1.23 - 2.14)*** | 0.95 (0.73 - 1.25) |
| Household owns a radio or television | 0.72 (0.57 - 0.91)** | 1.25 (0.99 - 1.58) | 1.16 (0.92 - 1.46) |
| Household wealth index tertile (ref. highest) |  |  |  |
| Lowest | 2.24 (1.59 - 3.16)*** | 0.66 (0.48 - 0.92)* | 0.39 (0.27 - 0.54)*** |
| Middle | 1.73 (1.33 - 2.24)*** | 0.87 (0.67 - 1.13) | 0.67 (0.52 - 0.86) ** |
| Number of shocks (ref. 0-1) |  |  |  |
| 2 | 1.54 (1.16 - 2.05)** | 0.99 (0.75 - 1.33) | 0.96 (0.71 - 1.29) |
| 3 | 1.48 (1.11 - 1.97)** | 1.23 (0.92 - 1.65) | 0.88 (0.65 - 1.19) |
| 4 | 2.18 (1.55 - 3.07)*** | 0.93 (0.67 - 1.30) | 0.82 (0.57 - 1.17) |
| Survey zone (ref. Central) |  |  |  |
| East | 0.63 (0.46 - 0.86)** | 1.64 (1.20 - 2.24)** | 0.56 (0.41 - 0.76)*** |
| North | 1.29 (0.91 - 1.82) | 0.82 (0.60 - 1.13) | 0.30 (0.21 - 0.44)*** |
| Livelihood zone (ref. Urban/peri-urban) |  |  |  |
| Pastoral | 0.87 (0.65 - 1.15) | 1.31 (0.99 - 1.74) | 0.74 (0.56 - 0.97)* |
| Agropastoral | 0.75 (0.58 - 0.98)* | 1.12 (0.86 - 1.46) | 1.06 (0.82 - 1.36) |
| Survey wave (ref. Wave 1) |  |  |  |
| Wave 2 | 0.79 (0.61 - 1.02) | 0.46 (0.35 - 0.61)*** | 1.19 (0.89 - 1.59) |
| Wave 3 | 0.82 (0.62 - 1.07) | 0.24 (0.18 - 0.31)*** | 1.46 (1.08 - 1.96)* |
| Wave 4 | 1.29 (0.97 - 1.72) | 0.19 (0.14 - 0.26)*** | 1.18 (0.87 - 1.61) |
| Wave 5 | 1.13 (0.84 - 1.54) | 0.14 (0.10 - 0.19)*** | 1.28 (0.92 - 1.77) |
| Wave 6 | 1.04 (0.76 - 1.42) | 0.20 (0.15 - 0.28)*** | 1.27 (0.91 - 1.77) |
| **Intermediate factors^b^** |  |  |  |
| Severe household food insecurity | -- | 0.56 (0.47 - 0.66)*** | 0.84 (0.70 - 1.01) |
| Caregiver consumes alcohol^c^ | -- | 1.45 (0.88 - 2.39) | 1.44 (0.83 - 2.50) |

^a^ Mutually adjusted for each other

^b^ Adjusted for the basic factors

^c^ N=3,160 observations, 583 children

*** p≤0.001, ** p≤0.01, * p<0.05

**Supplemental Table 2: Determinants of household food insecurity and child diet in Turkana**

|  | **Food insecurity** | **Child consumed**  **animal-source food** | **Child consumed**  **fruits or vegetables** |
| --- | --- | --- | --- |
|  | **aOR (95% CI)** | **aOR (95% CI)** | **aOR (95% CI)** |
| Number of children | **1204** | **1204** | **1204** |
| Number of observations | **6428** | **6428** | **6428** |
| **Basic factors^a^** |  |  |  |
| Female child (ref. male child) | 1.05 (0.88 - 1.25) | 1.00 (0.87 - 1.15) | 1.22 (1.04 - 1.43)* |
| Child aged ≥2 y (ref. <2 y) | 1.06 (0.87 - 1.30) | 0.73 (0.63 - 0.85)*** | 0.95 (0.79 - 1.15) |
| Caregiver educated (ref. not educated) | 0.72 (0.58 - 0.91)** | 1.49 (1.24 - 1.80)*** | 1.80 (1.47 - 2.19)*** |
| Caregiver age in y (ref. <25) |  |  |  |
| 25-34 | 0.58 (0.33 - 1.03) | 1.43 (0.94 - 2.18) | 1.06 (0.67 - 1.67) |
| 35 and above | 0.66 (0.36 - 1.22) | 1.55 (0.98 - 2.44) | 1.07 (0.65 - 1.75) |
| Caregiver marital status  (ref. married, only wife) |  |  |  |
| Unmarried | 1.01 (0.78 - 1.32) | 0.85 (0.70 - 1.04) | 1.11 (0.90 - 1.38) |
| Married co-wife | 0.91 (0.75 - 1.12) | 1.01 (0.86 - 1.19) | 0.94 (0.78 - 1.14) |
| Household head is female (ref. male) | 1.19 (0.97 - 1.45) | 0.94 (0.80 - 1.11) | 1.05 (0.87 - 1.26) |
| Caregiver is child’s biological mother | 0.97 (0.69 - 1.36) | 0.92 (0.72 - 1.18) | 0.91 (0.69 - 1.20) |
| Caregiver number of living children (ref. 0-2) |  |  |  |
| 3-5 | 1.30 (1.05 - 1.61)* | 0.64 (0.54 - 0.76)*** | 0.77 (0.63 - 0.93)** |
| 6 and above | 1.30 (0.97 - 1.74) | 0.63 (0.50 - 0.79)*** | 0.97 (0.75 - 1.26) |
| Household owns poultry | 0.59 (0.46 - 0.74)*** | 1.30 (1.09 - 1.55)** | 1.04 (0.85 - 1.26) |
| Household owns a radio or television | 0.95 (0.73 - 1.23) | 1.71 (1.38 - 2.11)*** | 1.44 (1.16 - 1.79)*** |
| Household wealth index tertile (ref. highest) |  |  |  |
| Lowest | 2.07 (1.53 - 2.82)*** | 0.63 (0.50 - 0.79)*** | 0.28 (0.22 - 0.37)*** |
| Middle | 1.57 (1.22 - 2.03)*** | 0.69 (0.57 - 0.84)*** | 0.43 (0.35 - 0.54)*** |
| Number of shocks (ref. 0-1) |  |  |  |
| 2 | 1.76 (1.30 - 2.40)*** | 0.96 (0.73 - 1.26) | 0.77 (0.58 - 1.03) |
| 3 | 2.19 (1.60 - 3.00)*** | 1.30 (0.99 - 1.70) | 0.79 (0.59 - 1.05) |
| 4 | 2.91 (2.06 - 4.11)*** | 1.30 (0.98 - 1.73) | 0.86 (0.63 - 1.17) |
| Survey zone (ref. Central) |  |  |  |
| North | 1.97 (1.46 - 2.67)*** | 1.06 (0.83 - 1.35) | 0.21 (0.13 - 0.32)*** |
| South | 2.22 (1.76 - 2.80)*** | 1.03 (0.84 - 1.25) | 0.98 (0.79 - 1.21) |
| West | 2.74 (2.11 - 3.56)*** | 1.10 (0.90 - 1.35) | 0.60 (0.47 - 0.76)*** |
| Livelihood zone (ref. Urban/peri-urban) |  |  |  |
| Pastoral | 1.27 (1.01 - 1.60)* | 1.11 (0.94 - 1.31) | 0.59 (0.49 - 0.72)*** |
| Agropastoral | 0.91 (0.68 - 1.22) | 1.20 (0.96 - 1.50) | 1.15 (0.91 - 1.45) |
| Fisherfolk | 1.10 (0.80 - 1.51) | 3.17 (2.44 - 4.13)*** | 0.32 (0.22 - 0.47)*** |
| Survey wave (ref. Wave 1) |  |  |  |
| Wave 2 | 1.09 (0.85 - 1.40) | 0.65 (0.55 - 0.78)*** | 0.64 (0.50 - 0.80)*** |
| Wave 3 | 1.53 (1.16 - 2.03)** | 0.40 (0.33 - 0.49)*** | 0.98 (0.78 - 1.24) |
| Wave 4 | 1.09 (0.83 - 1.43) | 0.30 (0.24 - 0.37)*** | 0.80 (0.63 - 1.03) |
| Wave 5 | 2.80 (2.01 - 3.90)*** | 0.20 (0.15 - 0.25)*** | 0.55 (0.42 - 0.73)*** |
| Wave 6 | 5.41 (3.67 - 8.00)*** | 0.36 (0.29 - 0.45)*** | 0.86 (0.66 - 1.11) |
| **Intermediate factors ^b^** |  |  |  |
| Severe household food insecurity | -- | 0.74 (0.62 - 0.89)** | 0.69 (0.57 - 0.85)*** |
| Caregiver consumes alcohol^c^ | -- | 1.19 (0.95 - 1.49) | 1.15 (0.89 - 1.50) |

^a^ Mutually adjusted for each other

^b^ Adjusted for the basic factors

^c^ N=6,423 observations, 1,203 children

***p≤0.001, ** p≤0.01, * p<0.05
